# Automatic Gender Detection in Twitter Profiles for Health-related Cohort Studies

**DOI:** 10.1101/2021.01.06.21249350

**Authors:** Yuan-Chi Yang, Mohammed Ali Al-Garadi, Jennifer S. Love, Jeanmarie Perrone, Abeed Sarker

## Abstract

**Objective:** Biomedical research involving social media (SM) data is gradually moving from population-level to targeted, cohort-level data analysis. Though crucial for biomedical studies, SM user’s demographic information (*e.g*., gender) is often not explicitly known from profiles. Here we present an automatic gender classification system for SM and we illustrate how gender information can be incorporated into a SM-based health-related study.

**Materials and Methods:** We used two large Twitter datasets: (i) public, gender-labeled users (Dataset-1), and (ii) users who have self-reported nonmedical use of prescription medications (Dataset-2). Dataset-1 was used to train and evaluate the gender detection pipeline. We experimented with machine-learning algorithms including support vector machines (SVMs) and deep-learning models, and released packages including M3. We considered user’s information including profile and tweets for classification. We also developed a meta-classifier ensemble that strategically uses the predicted scores from the classifiers. We applied the best-performing pipeline to Dataset-2 to assess the system’s utility.

**Results and Discussion:** We collected 67,181 and 176,683 users for Dataset-1 and Dataset-2, respectively. A meta-classifier involving SVM and M3 performed the best (Dataset-1 accuracy: 94.4% [95%-CI: 94.0%-94.8%]; Dataset-2: 94.4% [95%-CI: 92.0%-96.6%]. Including automatically-classified information in the analyses of Dataset-2 revealed gender-specific trends— proportions of females closely resemble data from the National Survey of Drug Use and Health 2018 (tranquilizers: 0.50 vs. 0.50; stimulants: 0.50 vs. 0.45), and the overdose Emergency Room Visit due to Opioids by CDC (pain relievers: 0.38 vs. 0.37).

**Conclusion:** Our publicly-available, automated gender detection pipeline may aid cohort-specific social media data analyses (https://bitbucket.org/sarkerlab/gender-detection-for-public).

## BACKGROUND

Social media data is increasingly being used for health-related research because of the large volume of salient information available from it. Users often discuss personal experiences or opinions regarding a variety of health topics, such as health services or medications. Such information can be categorized, aggregated and analyzed to obtain population-level insights,^1-5^ at low cost and in close to real time. It has thus been used as a resource for population health tasks such as influenza surveillance, pharmacovigilance and toxicovigilance.^6-8^ While early research mostly attempted to conduct observational studies on entire populations (*eg*., Twitter users discussing flu),^9^ some recent studies have been moving to targeted cohorts (*eg*., pregnant women,^10^ people in certain geo-locations,^11^ cancer patients,^12^ and people suffering mental health issues^13-16^). Demographic information about such cohorts can help researchers investigate what roles demographics have in a given study, understand if social media is biased towards specific cohorts, and explicitly address these biases.^17,18^ Funding agencies, including the National Institutes of Health (NIH), have emphasized the need to describe sex/gender information of the cohorts included in research studies (*eg*., through inclusion of women).^19^ This, however, presents a challenge for social media-based studies because the demographic information of the users are often not explicitly known.

One solution is to infer the demographic information from the user’s metadata. In the past two decades, researchers have developed various automatic methods for profiling users. Taking gender detection on Twitter as an example, researchers have investigated classification schemes based on the users’ (screen) names, profile descriptions, tweets, profile colors, and even images, with machine learning algorithms such as Support Vector Machine (SVM), Naive Bayes (NB), Decision Tree (DT), Deep Neural Network (DNN), and Bidirectional Encoder Representations from Transformers (BERT).^20-29^ However, only a few of them make their pipelines publicly available and have since been applied to social media mining tasks. For example, Sap et al.^24^ released a lexicon for gender and age detection and it was applied for mental health research.^13-15^ Knowles et al.^25^ released a package named Demographer to infer gender based on user’s first name and it was then employed to infer gender in studies for influenza vaccination^30^ and mental health.^16^ Wang et al.^31^ also released a multimodal deep learning system (M3) to infer gender based on user’s profile information, including picture, (screen) name, and description. Though these pipelines are available, we note that they have not been widely adopted by biomedical informatics researchers; possibly because not many researchers are aware of their existence, are concerned about the decrease of pipelines’ performances because of domain shift,^32^ or are even dubious on the validity of gender inference with machine-learning techniques. We also note that, to the best of our knowledge, the performances and utilities of these pipelines have not been evaluated using the targeted/domain-specific datasets.

Motivated by the above, we experimented with various strategies for developing a high-accuracy, automatic gender classification system using annotated datasets of general Twitter users, whose posts were retrieved in early 2020. We used held-out subsets of the gold standards to evaluate the performances of several classification strategies based on F_1_ scores and accuracies. Focusing on Toxicovigilance, we assessed the utility of the pipeline on a Twitter cohort of self-reported nonmedical consumers of prescription medications (PMs). Specifically, we applied the pipeline to infer the user’s gender and compared the observed gender distributions from Twitter to relevant metrics reported in other sources, namely the National Survey of Drug Use and Health (NSDUH) surveys^33^ and the CDC Wonder database.^34,35^ In this paper, we describe the development and evaluation of our automatic gender classifier on two datasets, and we illustrate the utility of our approach for deriving gender-specific insights from social media data. The source code for all experiments described will be made open source (https://bitbucket.org/sarkerlab/gender-detection-for-public).

## MATERIALS AND METHODS

This study was approved by the Emory University institutional review board (IRB00114235).

### Data collection

We collected publicly-available Twitter data from two sources: (i) the gender-labeled datasets (including user’s ID and gender label) for general Twitter users from previous work (Dataset-1), and (ii) the users who have self-reported nonmedical use of PMs (Dataset-2).

### Dataset-1: General Twitter Users

We collected gender-labeled datasets from Twitter, released by previous work.^22,23^ These datasets are constructed from Twitter users in general (ie., with no specific filtering criteria except for annotation) for the purpose of developing automatic gender detection scheme. The data from Liu & Ruths^22^ consists of 12,681 users with binary gender annotated via crowdsourcing through Amazon Mechanical Turk.^36^ The data from Volkova et al.^23^ consists of 800,000 tweets, randomly sampled from the data in Burger et al.^20^, which is labeled using user’s self-specified gender on Facebook or MySpace profiles linked to their Twitter accounts. Both datasets provide the users’ IDs and gender labels. Using twitter’s API, we extracted users’ publicly available data, including profile meta-data, such as handle names, descriptions, and profile colors, as well as the users’ timelines (only English tweets were collected while the retweets were excluded; users who had no original English tweets were dropped). We then combined the two datasets and split it into training (60%), validation (20%), and test (20%) sets for developing the pipeline.

### Dataset-2: Toxicovigilance

To conduct Toxicovigilance research using social media, we have collected publicly available, English tweets mentioning over 20 PMs that have the potential for nonmedical use or misuse (*eg*., Adderall®, Xanax®, and OxyContin®), from March 6, 2018 to January 14, 2020. We then developed automatic classification schemes to further detect the tweets that are describing self-reported nonmedical use (ie., misuse/abuse).^37,38^ Using a transformer-based classifier (fine-tuned from RoBERTa-large),^39^ which obtained the best performance on a gold standard, we classified all our collected tweets about prescription medications. For the tweets identified as self-report misuse/abuse, we extracted the users’ publicly available data, as in Dataset-1. In total, we obtained 176,683 users, who have at least one self-reported misuse/abuse of PMs. We were particularly interested in obtaining insights from social media on how gender plays a role in nonmedical PM use.

### Classification

Our strategy was to develop a gender detection classifier for each attribute in the user data (*eg*. name or tweets), which were treated as subtasks, and then constructed meta-classifiers based on the predicted scores from the aforementioned classifiers. An overview of the experiments is shown in Figure 1. Because each user’s data consists of a broad range of attributes, constructing a single classifier integrating all the derived features from the attributes might lead to inefficient model training and overfitting on a specific feature set, and thus it may be suboptimal. Our intuition was that combining the predicted scores of optimal classifiers on individual feature sets might increase model training efficiency, avoid overfitting, and provide an architecture that enables us to incorporate and iterate over different classifiers on each subtask.

**Figure 1:**
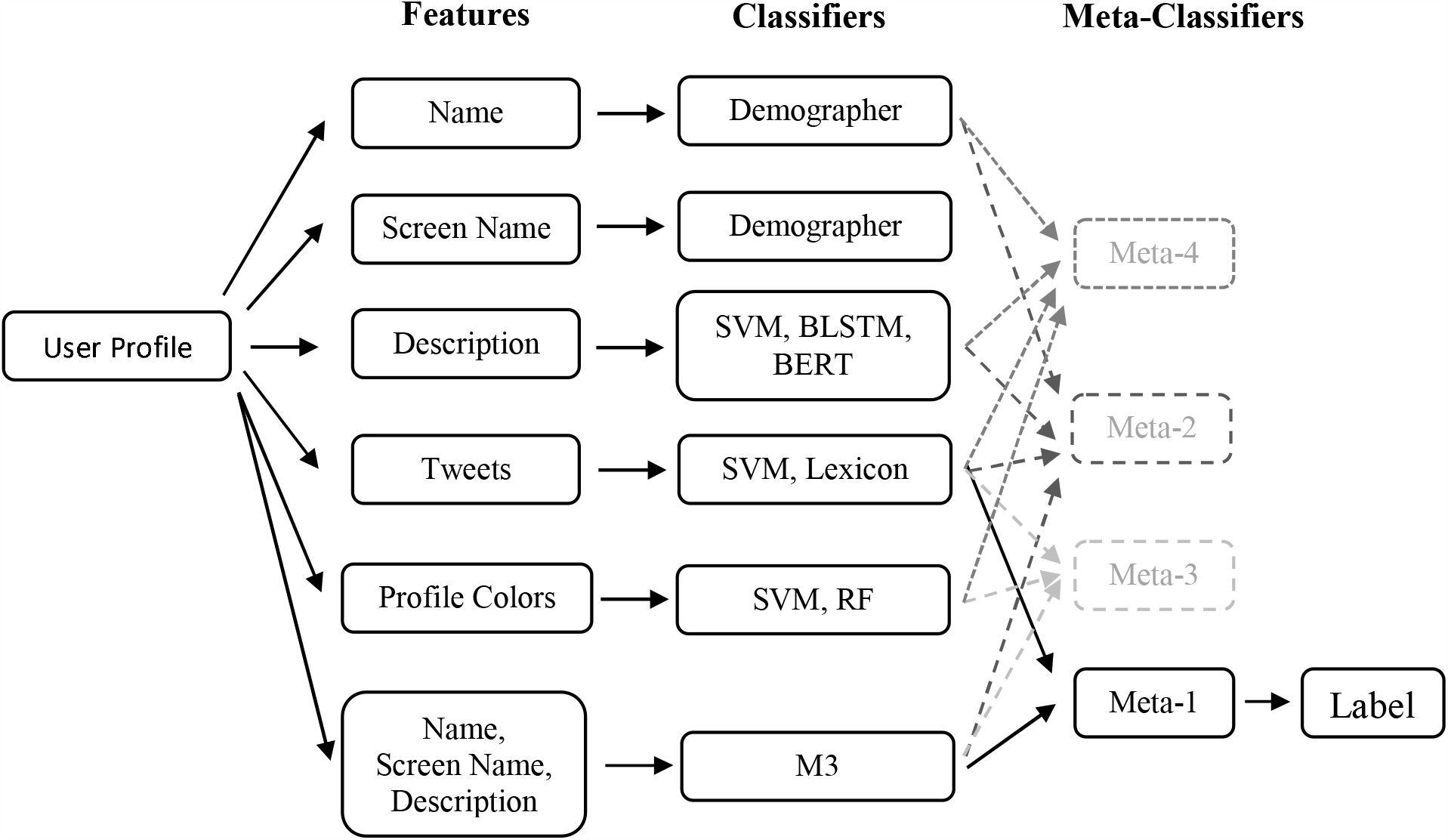
Gender classification pipeline, from user profile to gender label

We experimented with user’s attributes including name and screen name, description, tweets, and profile colors, while we considered the machine learning algorithms including SVMs,^40,41^ Random Forest (RF),^42^ bi-directional long short-term memory (BLSTM),^43,44^ and Bidirectional Encoder Representations from Transformers (BERT).^39,45^ We also experimented with the lexica released by Sap et al.^24^, the Demographer system by Knowles et al.^25^ and the M3 system (without profile picture) by Wang et al.^31^

The feature extraction and classification training for SVM and RF is done using the “Scikit-learn” package,^46^ the BLSTM classification is implemented using “Keras” package,^47^ and the BERT classification is implemented using package “simpletransformers” which is based on the package “transformers.”^48^ The details and hyperparameters are presented in the Supplementary Materials, Table S1.

### Name and Screen Name

We applied package Demographer^25^ (DG), version 1.0.4, on the users’ names. DG attempts to identify gender using character n-grams of user’s first name, trained using the list of given names from US Social Security data. Similar to DG, we trained a SVM classifier for screen names using character n-grams (n from 1 to 5).

## Description

To classify gender using a user’s description (ie., the bio text on each profile), we experimented with SVM, BLSTM, and BERT, approaches that are suited for free text data. BERT is a transformer-based model that produces contextual vector representations of words and achieves state-of-the-art performance on many tasks.^49,50^ Many models with similar architecture have then been implemented and released.^51,52^

Each description was pre-processed by lowercasing and anonymizing URLs and user names. For SVM, the features are the normalized term frequency of the 20,000 most frequent unigrams. For BLSTM and BERT, each word or character sequence was replaced with a dense vector, and the vectors were then fed into the relevant algorithms for training. We used Twitter GloVe word embeddings for the BLSTM^53^ classifier, where each word is converted to 200-dimensional vector. BLSTM was then trained with 20 epochs and dropout regularization and the best model was selected through accuracy on validation data. We chose to fine-tune RoBERTa-large for BERT algorithms.^39^ We trained the models with 1, 2, and 3 epochs and found that the model trained with 2 epochs performed the best.

### Tweets

For each user in the training data with at least 100 tweets, we merged all collected tweets as one single document as the training texts and experimented with SVMs. The pre-processing is the same as that for the SVM classifier using description. The regularization parameter was optimized according to the validation accuracy.

### Colors

We attempted to utilize five features associated with colors, which include profile background color, profile link color, profile sidebar border color, profile sidebar fill color, and profile text color. Each profile color is represented using RGB values, each value range from 0 to 255. To reduce the number of features for classification experiments, we divide each value into 4 groups, yielding 64 groups for each profile color. We then experimented with SVM and RF.

### Meta-classifier

We experimented with building meta-classifiers using a SVM classifier on the scores/probabilities estimated by the earlier classifiers. Our intuition is that combining different aspects learned by the classifiers may lead to a more thorough understanding of the data and, thus, better and more robust (low-variance) performance. Specifically, we experimented with four different combinations of the best classifiers on the given features:

- meta-1: SVM on tweets and M3 system.
- meta-2: SVM on tweets, M3 system, Demographer on name, and BERT on description.
- meta-3: SVM on tweets, M3 system, and SVM on colors.
- meta-4: DM on names, SVM on screen names, BERT on description, and SVM on tweets.

### Classification Performance Evaluation and Coverage

The classification performance evaluation is based on precision, recall, and F_1_ score (male and female separately), as well as accuracy (male and female combined). These metrics are defined as the follows:

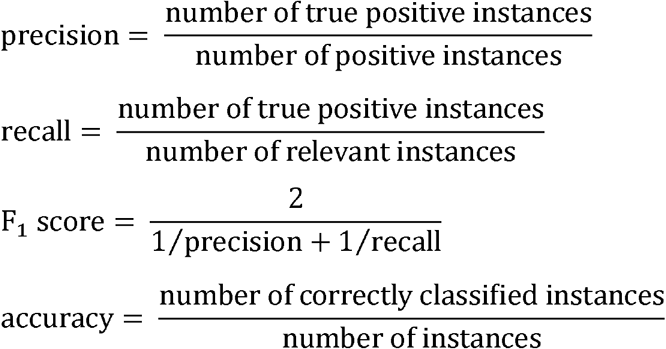

We also calculate the area under the receiver operating characteristic curve (AUROC). The ROC curve presents the relationship between the true positive rate and the false positive rate under different threshold and the AUROC provides a measure for the performance. The range of AUROC is from 0 to 1, with 1 being the best.

Some users have missing profile information such as name or description or use non-English characters in the name field. This may make the inference using the specific information impossible. Therefore, for each classifier, we show the percentage of users whose genders can be inferred from the relevant profile information (as “coverage”) while the performance is evaluated using this subset of users.

### Utility of the gender classifier: toxicovigilance

We applied the best-performing classification strategy on Dataset-2 and analyzed the classification results. Since Dataset-2 did not have manual binary annotations, we relied on a secondary source to identify a user’s gender—their self-identified gender information on the linked public Facebook profiles—whenever possible. The test set of Dataset-2 is composed of this subset of users.

We first focused on the test set to evaluate the performance of the classifier on this domain-specific dataset. We then analyzed the gender distribution of a large set of users who had self-reported misuse/abuse on one of the three abuse-prone PM categories— stimulants (eg., Adderall®), which can increase alertness, attention, and energy and are mostly prescribed to treat Attention deficit hyperactivity disorder (ADHD), tranquilizers (eg., alprazolam/Xanax®), which slow brain activity and are mostly used to treat anxiety, and pain relievers (eg., Oxycodone/OxyContin®), specifically for those containing opioids.^54,55^

We then compared the distributions with metrics from the 2018 NSDUH,^33^ conducted by the Substance Abuse and Mental Health Services Administration (SAMHSA),^55^ as well as the overdose-related Emergency Department Visits (EDV) in 2018 from the CDC Wonder database.^34,35^ We performed Pearson’s Chi-squared test to determine if the differences in female proportion inferred from different sources (gender classification on Twitter and survey) are statistically significant, defined as p-value < 0.05.

## RESULTS

### Data Collection

#### Dataset-1

In total, we were able to retrieve the user data from 67,181 users, consisting of 35,812 (53.3%) females (F) and 31,369 (46.7%) males (M), which is close to the distribution estimated by Burger et al.^20^ and Heil & Piskorski^56^ (55% female and 45% male) but deviate from the distribution estimated by Liu & Ruths^22^ (65% female and 35% male). The distribution is presented in Table 1.

**Table 1:** Data distribution of Dataset-1, the Training, Validation and Test sets from Dataset-1, Dataset-2 and Test dataset from Dataset-2 (users whose gender information is available)

| Dataset | F | M | Total |
| --- | --- | --- | --- |
| Training (Dataset-1) | 21,521 | 18,788 | 40,309 |
| Validation (Dataset-1) | 7,133 | 6,303 | 13,436 |
| Test (Dataset-1) | 7,158 | 6,278 | 13,436 |
| Total (Dataset-1) | 35,812 | 31,369 | 67,181 |
| Test (Dataset-2, with gender information available) | 155 | 258 | 413 |
| Total (Dataset-2) | - | - | 176,683 |

#### Dataset-2

We were able to retrieve past data from 176,683 users. Less than 0.3% of the users (413) had publicly available gender information from linked Facebook profile pages. 155 out of 413 users in this subset were female (37.5%), while 258 users were male (62.5%). This difference in male-female proportions is probably due to the differences in selection criteria: Dataset-1 used no selection criteria, while this subset required that the users need to have self-reported PM misuse/abuse on Twitter (as identified by our classifier).^38^

#### Dataset-1

The performance (F_1_-score, accuracy, and AUROC) for each classifier and meta-classifier are presented in Table 2, while the precisions and recalls are presented in the Supplementary Materials, Table S2.

**Table 2:**
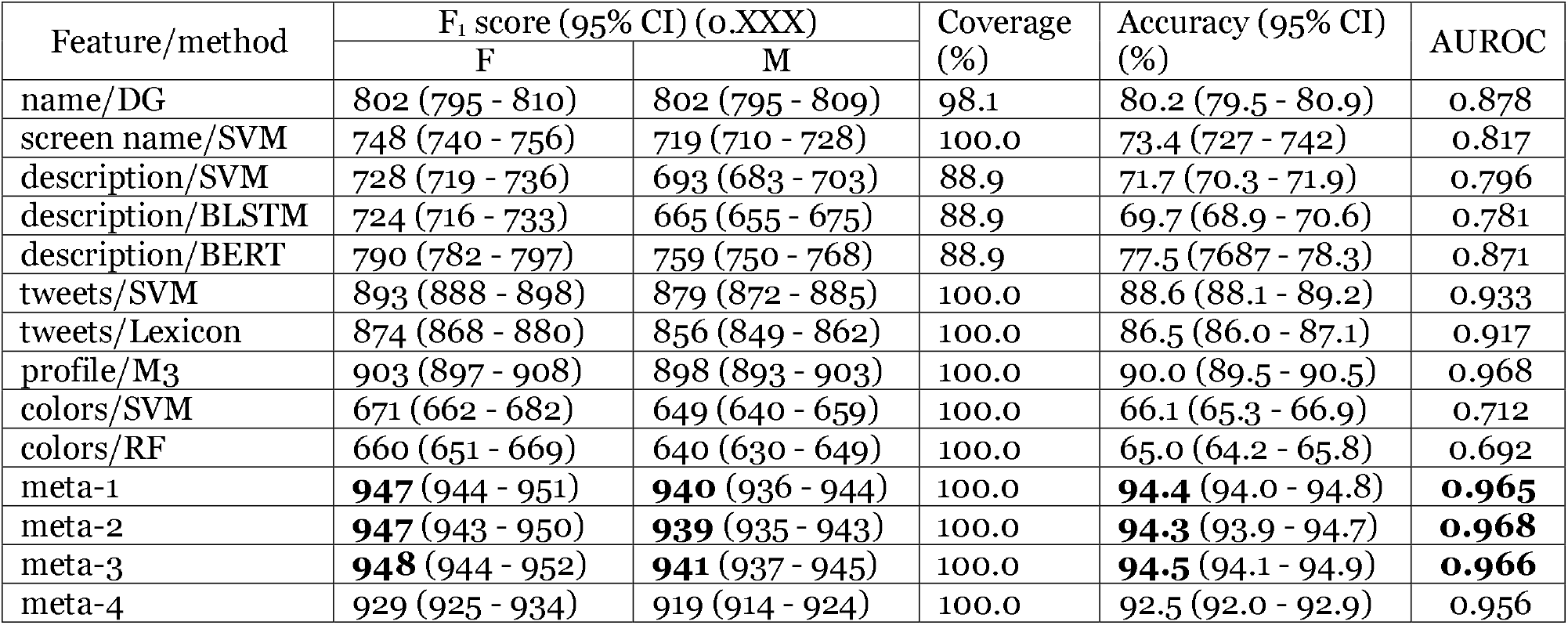
Test results (on Dataset-1) for classifiers, each based on different feature and/or method, and for meta-classifiers

### Name and Screen Name

The Demographer classifier has an accuracy of 80.2% and AUROC of 0.878, while the SVM classifier based on screen name has an accuracy of 73.4% and AUROC of 0.817. This shows that name is more informative than screen name for determining gender.

### Description

The best classifier on description was based on the BERT architecture, with an accuracy of 77.5% and an AUROC of 0.871, which is 6% higher in accuracy and 0.07 higher in AUROC than the SVM and BLSTM classifiers. This indicates that, to detect gender using short, self-describing sentences such as description on Twitter, the algorithms based on n-grams or context-independent vectors expression might not be enough; the algorithms based on contextual vectors performed better. All these classifiers performed worse than the Demographer applied on names, revealing that the names are more informative than the user’s description, in the context of gender detection.

### Tweets

The SVM classifier on tweets performed the second best among all classifiers tested, with an accuracy of 88.6% and an AUROC of 0.933, outperforming the lexica by Sap et al.,^24^ which was trained using similar pre-processing and method. The classifier’s performance depends on the amount of tweets available. In Figure 2, we plotted the accuracy versus the number of tweets. We found that the SVM’s accuracy can be increased to 90% if limiting the threshold to 400 tweets.

**Figure 2:**
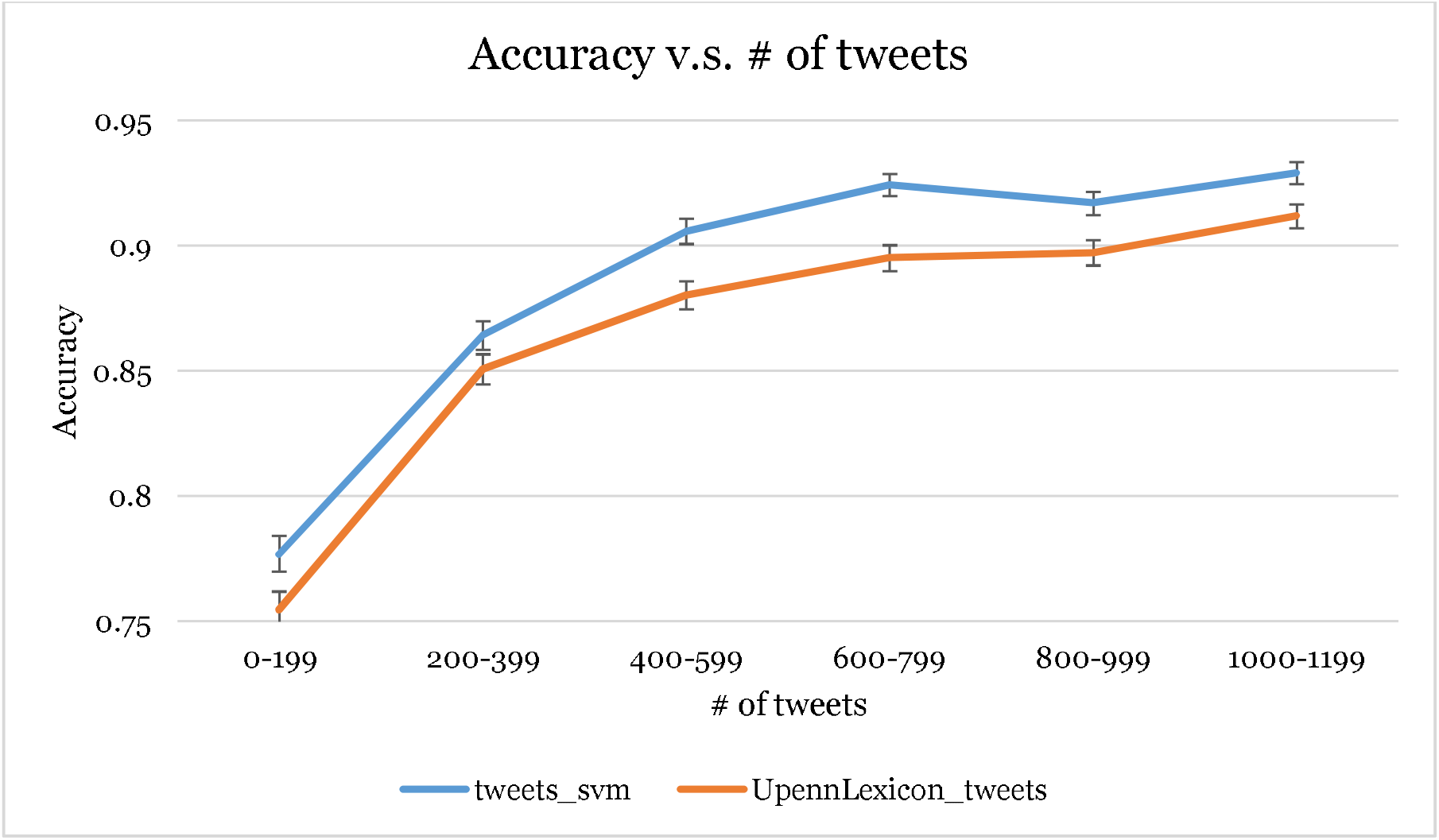
The SVM and Lexica’s performance’s dependence on the amount of tweets. The error bars mark 95% CI.

### M3 system

The M3 system by Wang et al.^31^ performed the best among all individual (non-ensemble) classifiers tested, with an accuracy of 90.0% and an AUROC of 0.968. This high performance suggests that using the collective features of name, screenname, and description can efficiently and accurately detect gender, even without the tweets.

### Colors

The classifiers using colors as features only produced accuracies around 66% (which is comparable to Alowibdi et al.^21^) and an AUROC of about 0.70. Though identifying gender using color is still possible to certain extent, it may not be suitable for biomedical research.

### Meta-Classifier

The first three meta-classifiers (meta-1 to −3) show similar performances, with an accuracy above 94%. Though meta-2 and meta-3 include the information learned by Demographer on name, BERT on description, or SVM on colors on top of M3 and SVM on tweets, they only show similar performances as meta-1, suggesting including these classifiers do not provide improvement. On the other hand, meta-4 yield only a lightly lower accuracy (92.4%), indicating that a complicated pipeline incorporating multiple users’ data as input only improves the performance by 2 points.

#### Dataset-2

The performance of the pipelines on the test set of Dataset-2 is shown on Table 3 (precisions and recalls are on Table S3 in the Supplementary materials). The best performing pipeline was meta-1 (accuracy 94.4%). Besides M3 and meta-1, all the classifiers experience performance drops possibly due to domain change. This supports that a more complicated deep learning pipeline on the user profile, such as M3, could be more robust than the classifiers trained on single user attributes (eg., BERT on description) or even the meta-classifier based on a number of such classifiers (eg., meta-4).

**Table 3:** Test results (on Dataset-2, for users who have revealed gender information on Facebook) for DG on name, BERT on description, SVM on tweets, Lexicon on tweets, M3, and meta-1

| Feature/Method | F<sub>1</sub> score (95% CI) (o.XXX) |  | Coverage (%) | Accuracy (95% CI) (%) | AUROC |
| --- | --- | --- | --- | --- | --- |
|  | F | M |  |  |  |
| name/DG | 722 (661 - 777) | 836 (799 - 870) | 0.95 | 79.3 (75.3 - 83.2) | 0.850 |
| screen name/SVM | 692 (633 - 745) | 777 (734 - 816) | 100.0 | 74.1 (697 - 782) | 0.839 |
| description/BERT | 672 (614 - 724) | 699 (646 - 748) | 0.95 | 68.6 (64.0 - 73.2) | 0.835 |
| tweets/SVM | 822 (773 - 866) | 894 (864 - 920) | 100.0 | 86.7 (83.3 - 89.8) | 0.909 |
| tweets/Lexicon | 775 (723 - 822) | 848 (813 - 880) | 100.0 | 81.8 (78.0 - 85.5) | 0.891 |
| profile/M3 | 894 (855 - 928) | 936 (913 - 957) | 100.0 | 92.0 (89.3 - 94.4) | **0.974** |
| meta-1 | **927** (894 - 954) | **955** (936 - 972) | 100.0 | **94.4** (92.0 - 96.6) | **0.965** |
| meta-4 | 882 (84.2 - 91.7) | 925 (899 - 947) | 100.0 | 90.8 (87.9 - 93.5) | 0.954 |

### Post-classification analyses

We applied meta-1 on all the users (176,683) and analyzed the gender distributions for the users who have self-reported abuse/misuse of three PM categories, tranquilizers, stimulants, and pain relievers (opioids). In Table 4, we report the number of users for each category, and the percentage of males and females, inferred through the classification results (meta-1), and reported by NSDUH 2018.^55^ For the NSDUH data, we calculated the male and female percentage from the tables that describe the estimated numbers of people who are at least 12 years old and have misused the specific medication categories in the past year, as surveyed in 2018 (Tranquilizer: Table 1.53A, Stimulants: Table 1.47A, Pain Relievers: Table 1.44A).

**Table 4:** Gender Distributions for Selected Medication Categories (inferred by the classifier / adjusted by the performance / and from NSDUH 2018)

| Medication Category | Number of Users | Percentage of Male / Female |  |  |
| --- | --- | --- | --- | --- |
|  |  | inferred | NSDUH 2018 | overdose EDV 2016 |
| Tranquilizers | 62,471 | 0.499/0.501 | 0.499/0.501 | - |
| Stimulants | 93,588 | 0.503/0.497<sup>†</sup> | 0.551/0.449 | - |
| Pain Relievers | 38,299 | 0.621/0.379<sup>†\$</sup> | 0.518/0.482<sup>*\$</sup> | 0.630/0.370 |

\* According to the Appendix A: Glossary, “Although the specific pain relievers listed above are classified as opioids, use or misuse of any other pain reliever could include prescription pain relievers that are not opioids. For misuse in the past year or past month, estimates could include small numbers of respondents whose only misuse involved other drugs that are not opioids.”
† The female proportion whose difference with the corresponding female proportion in NSDUH 2018 is statistically significant
\$ The female proportion whose difference with the corresponding female proportion in overdose EDV due to opioids in 2018 (CDC Wonder database) is statistically significant

For tranquilizer and stimulants users, the gender proportions inferred from Twitter are very close to the comparator from NSDUH 2018 (with no statistically significant difference for tranquilizer users), supporting the application of gender detection tools on social media data. In contrast, the gender proportion of pain reliever users is quite different from the comparator from NSDUH 2018, but much closer to the overdose EDV from CDC Wonder database.^34,35^ This suggests that Twitter data could be an indicator of the gender distribution of opioid overdoses and might provide complementary information to better understand the discrepancies between the two traditional data sources (ie., NSDUH and CDC Wonder).

## DISCUSSION

### Model Performance and Improvement

Our gender detection pipeline based on M3 system and tweets (meta-1) performs with high accuracy on both Dataset-1 (94.4%) and Dataset-2 (94.4%), while other methods (except M3) experience accuracy drops. This illustrates the importance of testing pipelines on the domain data of interest, and also provides clues for improving our pipeline. In the future, we plan to annotate part of Dataset-2 and incorporate it into the training/testing phase of the pipeline.

Besides incorporating Dataset-2 into training, using other classification algorithms or changing the model architecture may also improve the performance. For example, applying BERT models on tweets might improve the performance with the help of contextual embeddings. Incorporating multiple features in one system, similar to the M3 system,^31^ might further improve the performance. We chose our architecture based on model simplicity and robustness, and development efficiency, leaving investigation for further improvements to future work. The observed performances are also the highest reported in the literature to date.

### Toxicovigilance

Our post-classification analyses of the PM cohort illustrated the utility of automatic gender classification on social media data. The similarity of the gender proportions of tranquilizer and stimulant misusers from Twitter and those from NSDUH 2018 supports the effectiveness applying social media mining on Toxicovigilance. The inferred gender proportion of pain reliever users, though different from NSDUH 2018, is close to that of the overdose EDV according to the CDC Wonder database. This association between self-reports of drug use on Twitter and overdose EDV rates is consistent with our past research, in which we identified significant associations between opioid misuse reports on Twitter and overdose deaths over specific geolocations (*e.g*., counties and sub-states).^11^ These suggest that social media mining can provide insights on how the pain reliever misusers become victims of overdose and may even serve as an early warning system, if leveraged effectively. Social media provides the opportunity to combine multiple types of information about a single user, including past tweets, social connections, and geolocation. All the information combined can provide geolocation-, gender- and time-specific trends to extract insights and potentially test hypotheses such as the association between mental health issues and PM misuses. Furthermore, the surveillance can be done close to real time—not only a great improvement over the turnaround time for curating the overdose statistics and the NSDUH, but also enabling the possibility of intervention. For example, the system can provide treatment information to pregnant women who might be at risk of PM misuse, or people who have several risk factors of overdose. Note that we do not suggest that social media data analytics can replace the traditional resources, but that it may provide excellent complementary data if aggregated and curated accurately, and opportunity to provide information/intervention beyond the traditional health services.

### Limitations

The performance evaluation in Dataset-2 is limited by filtering criteria. The test data is biased toward the users who have linked their Facebook account to Twitter and provide their gender publicly. It is also subject to false information by the users—a problem that all survey-based studies have. Though this evaluation method provides an estimate without manual annotation, it is still better to annotate the gender on a random sample of Dataset-2 to evaluate the performance. The inference of the gender distribution for the PM misusers is also the PM abuse classification pipeline.^38^

The annotation methods also make the dataset subject to the bias introduced by the methods. The Dataset-1 is labeled in two ways: (1) the users’ Twitter profile picture, and (2) the user’s self-reported gender on Facebook or MySpace profiles. These render Dataset-1 being biased toward those whose gender can be identified, making it an approximation of the general Twitter users. Also, the gender inferred by the classifier is defined by the annotation methods, not strictly the same as the users’ gender identity or biological sex. The first (profile picture) may be closer to the user’s biological sex, while the other (self-reported gender) closer to gender identity. However, as it is estimated that less than 0.5% of the US population are considered as transgender (ie., a person whose gender identity is different than the biological sex),^57^ their contributions to the classification performance should not be significant in this work. We leave building a more comprehensive, non-binary classification scheme to future work.

### Ethics

Ethically speaking, though we only use the publicly available data and adhere to Twitter API’s use terms, we agree that Twitter users’ perception toward user profiling may vary.^58^ To avoid potential harms to the users, we only study and report on the aggregated data, not individual users. We also agree with the guidelines proposed in Williams et al.^58^ and make the pipeline publicly available to ensure reproducibility and transparency to researchers and Twitter users.

## CONCLUSIONS

As social media based health research focus is moving from population-level to cohort-level, incorporating user demographic information is becoming more important. In this work, we developed a gender detection pipeline and evaluated its performance on a general dataset and a domain-specific dataset. Our proposed pipeline shows high accuracy even when applied on a health-specific dataset. We further showed that the pipeline can be used to infer the PM misusers’ gender distribution, which is consistent with the statistical data reported by NSDUH 2018 (stimulants and tranquilizers) and by CDC Wonder database (overdose EDV due to Opioids). With the much-needed growing attention on explicitly incorporating demographic information, such as gender and race/ethnicity, in research, it is crucial to be able to conduct aggregated gender-specific analyses of health-related social media data. Our pipeline is readily usable by social media researchers who need to infer users’ demographics from their data. We note that, besides gender, other demographic information, such as race or age are also important for research, and developing pipelines for these user profiling tasks and evaluate them on domain specific datasets are part of our planned future work.

## Data Availability

The source code for all experiments described will be made open source.

https://bitbucket.org/sarkerlab/gender-detection-for-public

## AUTHOR CONTRIBUTIONS

YY conducted and directed the machine learning experiments, evaluations and data analyses, with assistance from MAA and AS. AS provided supervision for various aspects of the study. JSL and JP provided toxicology domain expertise for interpreting the results. YY drafted the manuscript and all authors contributed to the final manuscript.

## ACKNOWLEDGEMENTS

The authors thank the support from the National Institute of Health and National Institute of Drug Abuse.

