## Supplementary Materials for "Automatic Gender Detection in Twitter Profiles for Health-related Cohort Studies"

Postal address: 101 Woodruff Circle, 4th Floor East, Atlanta, GA 30322

**Table S1:** The details and hyper-parameters for the classification experiments.

| Feature/Method | hyper-parameters and details |
|---|---|
| screen name/SVM | features: the counts of 10,000 most frequent character n-grams where n is from 0 to 5<br>Linear SVC with C=1 |
| description/SVM | features: the normalized term frequency of the 20,000 most frequent unigrams<br>tokenizer: happierfuntokenizing[1] with minor change for python 3<br>LinearSVC with C=1 |
| description/ BLSTM | Tokenization:<br>num_words: 30,000, maxlen: 128<br>Word Embeddings:<br>Twitter Glove, 200 dimensional vectors<br>Model:<br>an embedding layer [Output Shape: (None, 128, 200)] followed by a bidirectional LSTM layer [Output Shape: (None, 200)], followed by a dense layer [Output Shape: (None, 2)]<br>training:<br>loss function: 'categorical_crossentropy'<br>optimizer: 'adam'<br>dropout rate: 0.2<br>recurrent dropout: 0.2<br>metrics: accuracy<br>epochs: 20 |
| description/BERT | Tokenization:<br>max_seq_lenth: 128<br>Model:<br>RoBERTa-large<br>Training:<br>train_batch_size: 32<br>num_train_epochs: 2<br>learning_rate: 5e-6 |
| tweets/SVM | feature extraction and tokenizing are the same as description/SVM<br>LinearSVC with optimal C=32 |
| colors/SVM | LinearSVC with C=1 |
| colors/RF | n_estimators = 1000 |

**Table S2:** The precision and recall of the test results (on Dataset-1) for classifiers, each based on different feature and/or method, and for meta-classifiers

| Feature/method | precision | | recall | |
|---|---|---|---|---|
| | F | M | F | M |
| name/DG | 0.855 | 0.755 | 0.755 | 0.855 |
| screen name/SVM | 0.756 | 0.710 | 0.740 | 0.728 |
| description/SVM | 0.684 | 0.693 | 0.757 | 0.611 |
| description/BLSTM | 0.697 | 0.698 | 0.754 | 0.635 |
| description/BERT | 0.779 | 0.771 | 0.801 | 0.747 |
| tweets/SVM | 0.895 | 0.877 | 0.891 | 0.880 |
| tweets/Lexicon | 0.873 | 0.857 | 0.875 | 0.854 |
| profile/M3 | 0.941 | 0.861 | 0.867 | 0.938 |
| colors/SVM | 0.694 | 0.628 | 0.650 | 0.673 |
| colors/RF | 0.684 | 0.616 | 0.637 | 0.664 |
| meta-1 | 0.948 | 0.939 | 0.946 | 0.941 |
| meta-2 | 0.946 | 0.940 | 0.947 | 0.939 |
| meta-3 | 0.950 | 0.939 | 0.946 | 0.943 |
| meta-4 | 0.927 | 0.921 | 0.931 | 0.917 |

**Table S3:** The precision and recall of test results (on Dataset-2, for users who have revealed gender information on Facebook) for DG on name, BERT on description, SVM on tweets, Lexicon on tweets, M3, and meta-1

| Feature/ Method | precision | | recall | |
|---|---|---|---|---|
| | F | M | F | M |
| name/ DG | 0.724 | 0.834 | 0.719 | 0.837 |
| screen name/SVM | 0.625 | 0.842 | 0.774 | 0.721 |
| description/ BERT | 0.558 | 0.861 | 0.846 | 0.588 |
| tweets/ SVM | 0.825 | 0.892 | 0.819 | 0.895 |
| tweets/Lexicon | 0.725 | 0.889 | 0.832 | 0.810 |
| profile/M3 | 0.891 | 0.938 | 0.897 | 0.934 |
| meta-1 | 0.918 | 0.961 | 0.935 | 0.950 |
| meta-4 | 0.850 | 0.947 | 0.916 | 0.903 |

1. Sap M, Park G, Eichstaedt J, et al. Developing age and gender predictive lexica over social media. Paper presented at: Proceedings of the 2014 Conference on Empirical Methods in Natural Language Processing (EMNLP)2014.
